# Effect of energy shortages on institutional delivery in India

**DOI:** 10.1101/2022.11.16.22282404

**Authors:** Eugenia Amporfu, Bridget R. Irwin, Benjamin Sas Trakinsky, Karen A. Grépin

## Abstract

**Introduction:** Energy shortages are a common challenge in many low- and middle-income countries and can disrupt the operation of healthcare facilities, which can compromise patient outcomes and affect health service utilization. Maternal healthcare use, in particular, has been found to be negatively correlated with power outages in other contexts. The following study investigates the association between state-level energy shortages and institutional delivery rates in India and how the association varies according to women’s socio-economic status.

**Methods:** Using data from the 1998-99 and 2005-06 India Demographic and Health Surveys, along with information on power outages from India’s Central Electricity Authority, we estimate the association between energy shortages and institutional delivery rates using both logistic and multinomial regressions.

**Results:** Energy shortages were associated with reduced rates of institutional delivery: a 10% increase in the shortage level corresponded to a 1.1% decline in the percentage of women giving birth in a healthcare facility. Deliveries in public health facilities were more likely to be disrupted by energy shortages than deliveries in private facilities.

**Conclusion:** Energy shortages are an important determinant of institutional delivery in India. Given that increasing institutional delivery rates is likely important to reduce maternal mortality, policymakers should work to mitigate the impact of energy disruptions on healthcare seeking behaviours.

## Introduction

Energy shortages occur when the demand for electricity exceeds the supply, and have been linked to a number of negative outcomes, included reduced economic development due to financial and productivity losses (1), and poorer health (2,3). Indeed, electricity has been identified as an important driver of human development, and there is evidence to suggest that electrification, defined as connecting a place to a supply of electricity (3), can lead to reduced morbidity (4–6) and mortality (7–9), increased care-seeking (10,11), and improved quality of care (9). Unreliable electricity supply on the other hand, which is characterized by the inability of the electric system to supply the energy requirements of all consumers at all times or to withstand disturbances in system components (12,13), can severely impede healthcare access, delivery, and quality, disrupting the use of medical and diagnostic devices, as well as essential communication, transportation, and emergency services (14–16). Given these effects, expanding access to reliable electricity has become a global development priority and Target 7 of the United Nation’s Sustainable Development Goals (SDGs) calls for establishing “access to affordable, reliable, sustainable, and modern energy for all” (17). Despite this, access to electricity remains uneven and almost 1.1 billion people worldwide continued to lack an electricity connection in 2016, the majority of whom resided in rural and resource-constrained settings (18). As a large country with a population exceeding 1.3 billion, India accounts for approximately one quarter of all people without electricity access (18), and is plagued by widespread and frequent energy shortages (1). These shortages have been attributed to a lack of infrastructure and maintenance funding and investment due to a combination of low energy prices and inadequate government subsidies to distribution companies (1). Although progress has been made, with Indian Prime Minister Narendra Modi declaring all villages in the country electrified as of April 2018, fewer than 10% of these villages achieved 100% household connectivity, and power outages remain common (19).

Power outages can have devastating consequences, particularly in health facilities, and have been implicated in a number of high mortality events across the country (20–22). In 2016, for example, 21 patients died in a Telangana State hospital after a black out resulted in the failure of important life support systems, such as ventilators and incubators (20), while in 2015, a flood in Tamil Nadu knocked out the power supply to hospital ventilators, leading to 18 deaths (21). In addition to direct harm caused to patients, the frequent power outages that continue to occur across India have also been found to impact care-seeking behaviours, including among expectant mothers (23). Although the overall maternal mortality rate (MMR) in India has been steadily declining over the past several years; from 556 deaths per 100,000 live births in 1990 to 167 in 2013 (24); it still greatly exceeds the global target outlined in goal three of the SDGs of fewer than 70 deaths per 100,000 live births (25), and India accounts for approximately 20% of global maternal deaths (26). As a result, the government of India has promoted institutional delivery as a key strategy for improving maternal outcomes in India, encouraging women to give birth in a health facility in the presence of a skilled health professional wherever possible [17,20,21]. There is evidence to suggest, however, that power outages may undermine the benefits of institutional delivery, reducing a woman’s likelihood of seeking or receiving maternal healthcare (23).

Reductions in the quality of care provided in health facilities during an energy shortage, for example due to the inability to operate essential medical and diagnostic devices, provide lighting and refrigeration, or perform emergency procedures, may prevent a woman from receiving the care she needs in a timely manner (29,30), or may deter her from seeking care at a facility all together. Disruptions in transportation and communication systems may also inhibit a woman’s access to care (14,23), particularly in rural settings where the distance to the nearest health facility tends to be greater (29), while among women who do seek care at an institution, increased demand for health services during power outages may result in over-crowding, staff shortages, and heightened waiting times, increasing the likelihood of being turned away (23).

A recent systematic review summarizing the health impacts of electrification and electricity reliability in low- and middle-income countries (LMICs) found that gaining access to electricity was generally associated with an increased use of maternal health services, including antenatal care and family planning, as well as reduced maternal mortality (3). A cross-sectional study in India identified a greater odds of antenatal care use among rural women residing in electrified villages compared to those in unelectrified villages (11), while a longitudinal study reported similar positive associations between village electrification status and antenatal care visits in Pakistan (10). A significant negative correlation between maternal mortality rates and the percentage of households in the district with electricity was also observed in India (8), and a study in Uganda found that having electricity at a healthcare facility had a protective effect on maternal deaths, as a result of their increased capacity to provide emergency obstetric care (9). Along with maternal health more broadly, links between energy shortages and institutional delivery were also identified, and Köroglu et al. (2019) (23) found that a higher frequency of power outages was significantly associated with a lower odds of delivering in a health facility in Maharashtra State, India, while both Singh (2016) and Kumar et al. (2014) demonstrated that an inadequate supply of electricity in health facilities can lead to a significant reduction in institutional delivery rates (32,33). Furthermore, a qualitative analysis exploring barriers to institutional delivery in India identified a lack of electricity in health facilities as one of the factors that prevented women from giving birth in an institution, with respondents noting that the conditions at these facilities are no better than home (34).

While the importance of reliable electricity for maternal health and institutional delivery has been well-demonstrated, the distributional consequences of a lack of access to electricity by socio-economic status are less well understood. Poor people, for example, who have limited access to electricity in their everyday lives, as well as those living in rural areas where electricity is less widely available, may be less affected by energy shortages, and their healthcare utilization patterns may not change significantly. On the other hand, people in higher wealth quintiles and who reside in more urban areas may be dependent on electricity, and may therefore feel its loss more acutely, altering their healthcare seeking behaviours to account for anticipated declines in quality of care or delays in travel. Given these potential associations, the following paper seeks to investigate the impact of state-level energy shortages on institutional delivery rates in India, both overall and by income status and area of residence. The results will help to quantify the magnitude of the impact of energy shortages on institutional delivery rates in India, as well as to identify the women who are most vulnerable to power outages, allowing for the development of strategies to mitigate the impact of unreliable access to electricity on maternal health outcomes.

### Conceptual Framework

The nature of the problem at hand can be visualized in a form of hierarchical data. First, the individual decides whether to deliver in a health facility or in a home. The decision is affected by many factors including the woman’s characteristics such as age, wealth status, education, marital status, presence of complications, gender of household head and location of residence. Second, the decision is also affected by factors that are beyond the control of the woman, but as a result of public policy, and this includes the availability of electricity. Electricity supply is undertaken by the state in which the woman lives, implying that women who live in the same state would have similar experience with electricity supply. The third category is time. Women who deliver in a particular year could have some similar experiences due to the period which is affected by global, national, or state factors. Taking the hierarchy of data into account is important to ensure proper estimation.

## Materials and methods

### Data source

This study analyzes data from the 1998-99 and 2005-06 rounds of the Indian Demographic and Health Survey (DHS), a nationally representative survey of women aged 15 to 49 years that collects data on a number of health indicators, including family planning and the use of maternal and reproductive health services. Births that occurred less than 5 years preceding the survey were included in the study, and the final sample thus contains information on 77,583 births, spanning the years 1992 to 2006. To estimate energy shortages during this time period, we made use of state-level data from the Indian Energy Repository, which was extracted by Allcott et al. for their 2015 analysis of the effects of energy shortages in India on industry (1) and can be accessed at www.indiaenergydata.info. Using state level data on energy shortage is important as it is able to capture the variation in electricity supply across the states (41) Data were available for the years 1992 to 2011, with the exception of certain years, such as 1994-95 and 2003, for which data was missing for some states. Besides, the data on energy shortage does not exceed 2011, hence the DHS data 2015-16 was not included. Overall, the merged dataset containing information on both births and energy per state, per year was large, with 77,583 observations.

### Measures

Institutional delivery was assessed using self-reported data on the location of delivery of all births that occurred within 5 years preceding the date of the survey. Deliveries that took place in a public or private healthcare facility were defined as institutional, while those that took place at home or at another person’s home were considered non-institutional. For analysis purposes, institutional delivery was treated as a binary variable in logistic regressions, with categories institutional and non-institutional, and as a nominal variable in multinomial regressions, with 3 categories: non-institutional, public institution, and private institution.

Energy shortage was computed in accordance with Indian Central Electricity Authority (CEA) as described by Allcott et al.(1) using the following equation:

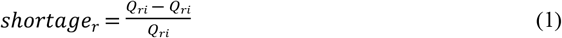

where *Q*_*di*_ represents the assessed demand for energy in the consumer’s state in a given year, and *Q*_*si*_ represents energy available in the same state and year. Thus shortage, *shortage*_*r*_, describes the extent to which supply lags demand for energy in a given state in a given year (1). For analysis purposes, energy shortage was treated as a continuous variable representing the percentage of demand not met, and ranged from approximately 23.8% in 1992-1993 to 10.8% in 1999-2002.

Additional data on the characteristics of the birth, mother, and household were also extracted from the DHS and were included in the analyses as covariates. Birth characteristics included birth month (nominal categorical variable) and birth order of the child, which was considered a proxy for parity (ordinal categorical variable). Mother characteristics included mother’s age at delivery (ordinal categorical variable), education (ordinal categorical variable), marital status (nominal categorical variable), religion (nominal categorical variable), and ethnicity (nominal categorical variable). Household characteristics included urbanicity (binary variable), state of residence (nominal categorical variable), gender of household head (binary variable), and wealth quintile (ordinal categorical variable). Additional data that was used for robustness checks included the number of antenatal care visits attended (continuous variable), and pregnancy complications (binary variable). In another robustness check, we also controlled for rain in the birth month which was obtained from Open Government Platform in India^2^.

### Data analysis

Statistical analyses were conducted using Stata 15.0. Descriptive statistics, including means and frequencies, were calculated to describe the characteristics of the sample, as well as energy shortages in India. The impact of electricity shortages on institutional delivery was modeled using the equation:

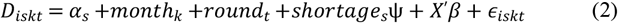

Where *D*_*is*_ is a dummy equaling one when woman *i* from state *s* delivered in an institutional facility (either private or public) in month *k* and round *t*, and zero otherwise, *α*_*s*_, *month*_*k*_, and *round*_*t*_ are fixed effects for the state of delivery, month of the delivery, and the round when the data was collected, respectively, shortage represents the unmet demand for electricity in the state *s* during the corresponding year of delivery, and *X* is a vector of control variables including birth order, maternal age, education, religion, ethnicity, and marital status, and household wealth quintile and urbanicity.

As already explained, the study combined repeated cross sectional micro data and time series macro data. The women who delivered, whether at home or in institutions, are nested in the states in which the lived, and the states are nested in time: month and year. Thus, classifying the variables in *X* as level one variables, state variables including energy shortage are level two variables; month of delivery as level three variable and year as level four variable. This creates four level hierarchical data which is suitable for multilevel modelling. Using the conventional regression methods of estimation would violate the assumption of constant variance and independent observations and hence yield unreliable results. The multilevel modelling takes the nested nature of the data into account to improve the efficiency of estimation. Since the outcome variable is binary, multilevel regression is appropriate. The four-level model can be specified below as:

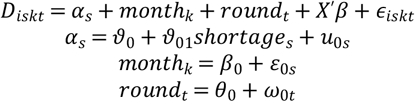

The first equation is the level one equation and has the variables at all levels as well as the level one or individual level variables *X*. The ‘fixed effects’: *α*_*s*_, *month*_*k*_, and *round*_*t*_ are modelled as random intercepts to allow for variation in each level unit. The state of delivery intercept, *α*_*s*_ is simultaneously estimated in the level two equation which is the second equation. Energy shortage, a level two variable, is used to predict the intercept which is modelled to account for variation in the chances of delivering in an institution across states. The two other random intercepts, *month*_*k*_ and *round*_*t*_, specified in the third and fourth equations, are for the levels three and four respectively. Each intercept measures the log odds of institutional delivery when all other variables equal zero, and all are specified as random intercept to account for variation in institutional deliveries due to state of delivery, month and the year of delivery.

The two common tests for the variability of the chances of institutional delivery across states, months, and rounds are the Intraclass Coefficient (ICC) and the likelihood ratio test. The ICC measures the proportion of variation in the chances of delivering in an institution that is explained by variability in institutional delivery between states, or between months or between rounds. While a zero ICC represents no variation across the level variables, implying no need for multilevel modelling, a high ICC represents high variation across level variables.

Both logistic regression, in which institutional delivery was treated as a binary variable and defined as institutional if a woman gave birth in a public or private health facility and non-institutional otherwise, and multinomial regression, in which institutional delivery was treated as a nominal categorical variable with home births defined as 0, public health facility births defined as 1, and private health facility births defined as 2, were performed. This was to account for previously demonstrated perceived differences in the quality of care delivered at public compared to private healthcare institutions, which tend to be better equipped (42), that may affect a woman’s healthcare seeking behaviours. In each regression, marginal effects were computed for the shortage variable to estimate the magnitude of the effect of energy shortages on institutional delivery. In an additional analysis, interaction terms were included to assess whether the effect of energy shortage on institutional delivery differs according to socioeconomic status and urbanicity. In addition, due to potential concerns about potential confounding effects from rain (rain may influence the likelihood of an energy shortage and the likelihood a woman gives birth in a clinic), we also test one specification in which we control for rain in our estimates. Adjusted results are presented. P-values less than 0.05 were considered statistically significant.

## Results

### Characteristics of the sample

Sample characteristics, including the characteristics of the births, mothers, and households included in the study, as well as information on energy shortages in India during the survey period, are presented in Table 1. A positive trend in both public and private institutional deliveries was observed over time, with institutional delivery rates increasing from approximately 26% in 1992-93 to over 46% in 2004-06. Correspondingly, the percentage of deliveries occurring at home decreased from nearly three quarters (74.0%) in 1992-93 to just over half (53.6%) in 2004-2006. The average number of antenatal care visits attended by a mother also increased during this period, from 2.67 visits to 3.86. Across all survey years, the majority of mothers were 20-34 years of age, and a significant minority had tertiary education, although the proportion with a secondary education or higher increased steadily over time, from 24.9% in 1992-93 to 46.8% in 2004-06. Over 60% of households were situated in rural locations regardless of the survey year, consistent with Indian data which shows the population of India is predominantly rural, and just a small proportion (6.3%-10.1%) were headed by a female. On average, the demand for energy exceeded the supply, and energy shortages ranged from 10.8% to 23.8% depending on the survey year.

**Table 1:**
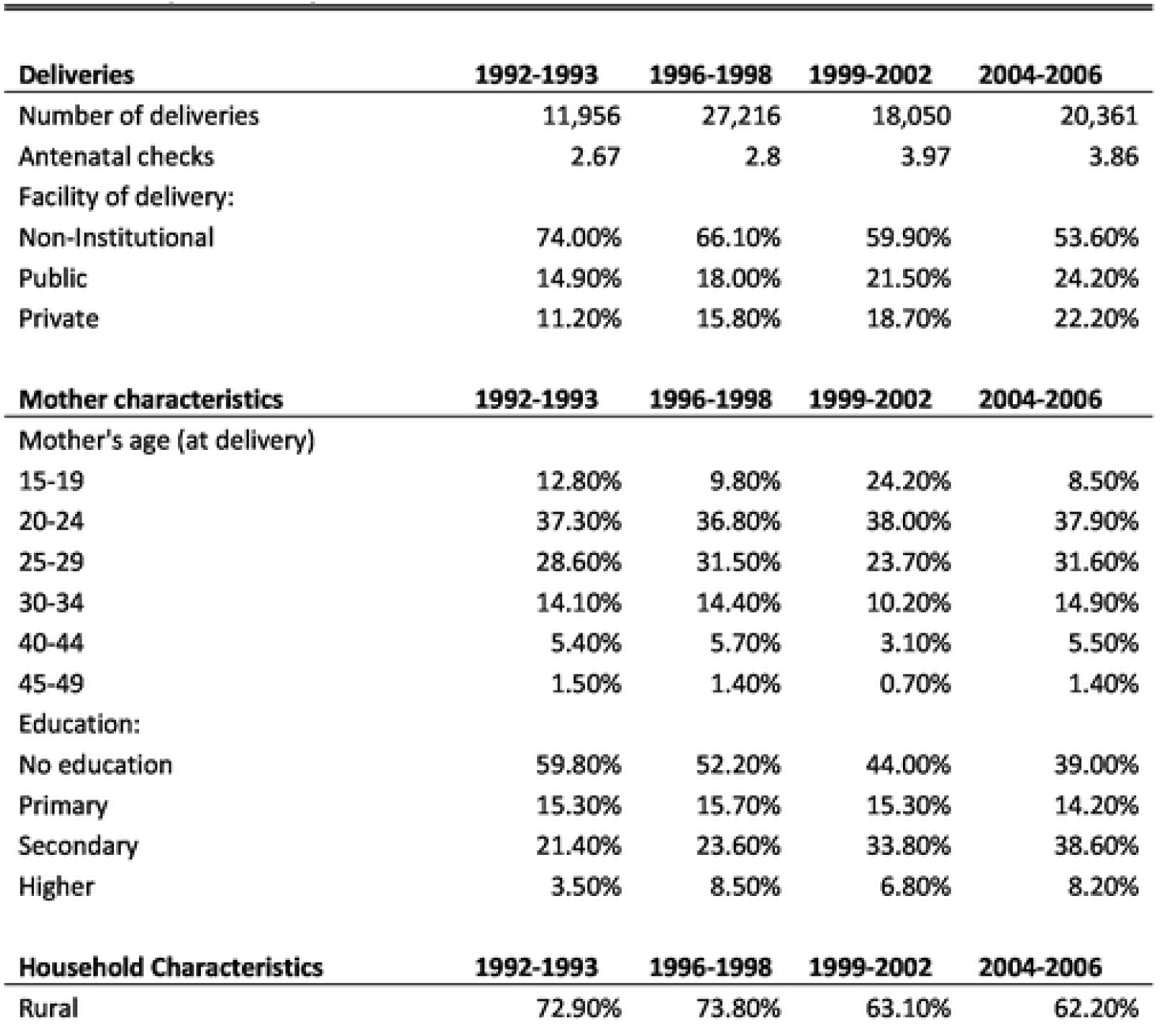
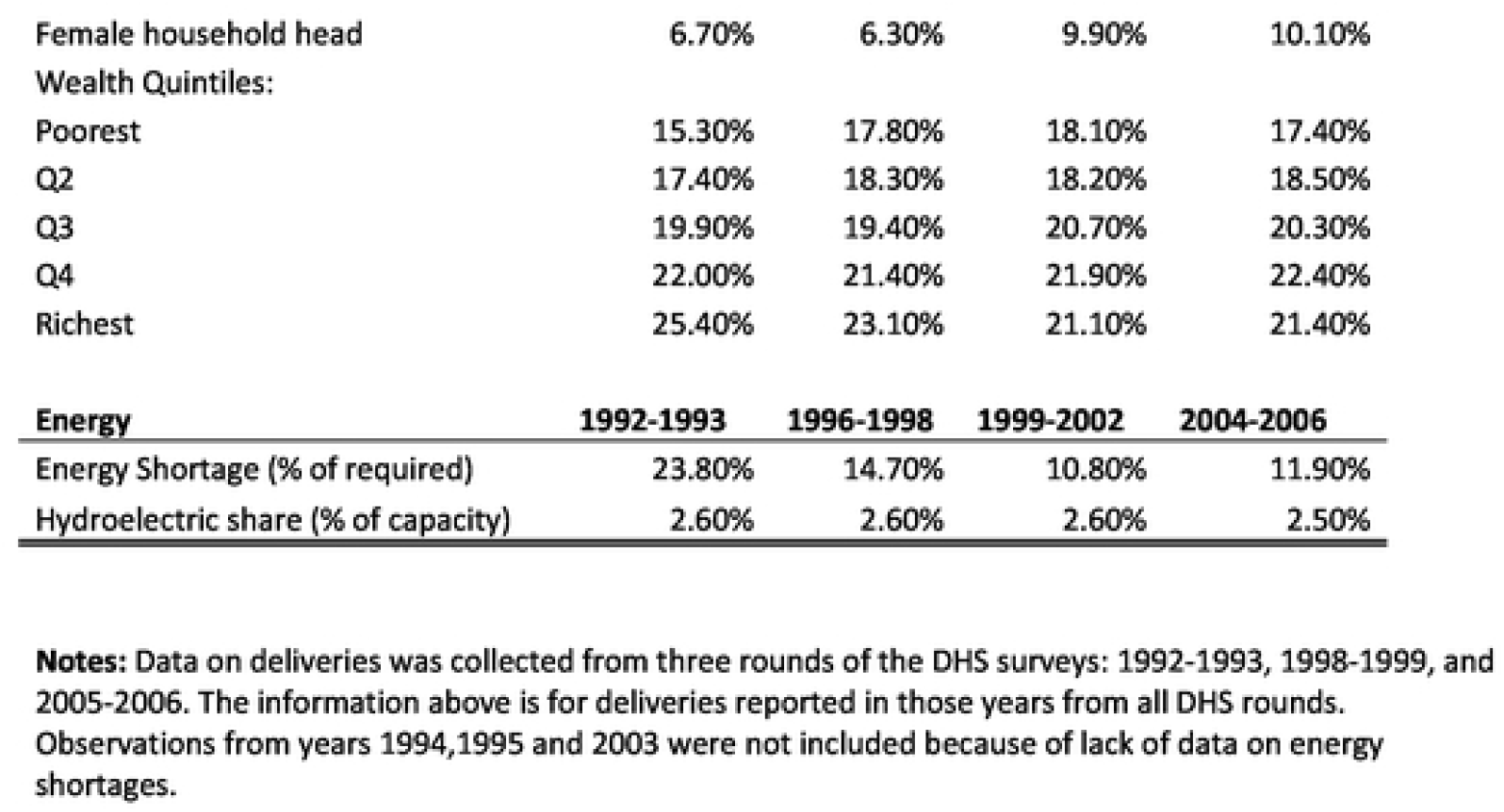
Sample summary statistics.

| **Deliveries** | **1992-1993** | **1996-1998** | **1999-2002** | **2004-2006** |
| --- | --- | --- | --- | --- |
| Number of deliveries | 11,956 | 27,216 | 18,050 | 20,361 |
| Antenatal checks | 2.67 | 2.8 | 3.97 | 3.86 |
| Facility of delivery: |  |  |  |  |
| Non-Institutional | 74.00% | 66.10% | 59.90% | 53.60% |
| Public | 14.90% | 18.00% | 21.50% | 24.20% |
| Private | 11.20% | 15.80% | 18.70% | 22.20% |
| **Mother characteristics** | **1992-1993** | **1996-1998** | **1999-2002** | **2004-2006** |
| Mother's age (at delivery) |  |  |  |  |
| 15-19 | 12.80% | 9.80% | 24.20% | 8.50% |
| 20-24 | 37.30% | 36.80% | 38.00% | 37.90% |
| 25-29 | 28.60% | 31.50% | 23.70% | 31.60% |
| 30-34 | 14.10% | 14.40% | 10.20% | 14.90% |
| 40-44 | 5.40% | 5.70% | 3.10% | 5.50% |
| 45-49 | 1.50% | 1.40% | 0.70% | 1.40% |
| Education: |  |  |  |  |
| No education | 59.80% | 52.20% | 44.00% | 39.00% |
| Primary | 15.30% | 15.70% | 15.30% | 14.20% |
| Secondary | 21.40% | 23.60% | 33.80% | 38.60% |
| Higher | 3.50% | 8.50% | 6.80% | 8.20% |
| **Household Characteristics** | **1992-1993** | **1996-1998** | **1999-2002** | **2004-2006** |
| Rural | 72.90% | 73.80% | 63.10% | 62.20% |
| Female household head | 6.70% | 6.30% | 9.90% | 10.10% |
| Wealth Quintiles: |  |  |  |  |
| Poorest | 15.30% | 17.80% | 18.10% | 17.40% |
| Q2 | 17.40% | 18.30% | 18.20% | 18.50% |
| Q3 | 19.90% | 19.40% | 20.70% | 20.30% |
| Q4 | 22.00% | 21.40% | 21.90% | 22.40% |
| Richest | 25.40% | 23.10% | 21.10% | 21.40% |

| Energy | 1992-1993 | 1996-1998 | 1999-2002 | 2004-2006 |
| --- | --- | --- | --- | --- |
| Energy Shortage (% of required) | 23.80% | 14.70% | 10.80% | 11.90% |
| Hydroelectric share (% of capacity) | 2.60% | 2.60% | 2.60% | 2.50% |

**Notes:** Data on deliveries was collected from three rounds of the DHS surveys: 1992-1993, 1998-1999, and 2005-2006. The information above is for deliveries reported in those years from all DHS rounds. Observations from years 1994,1995 and 2003 were not included because of lack of data on energy shortages.

### Prediction of institutional delivery by energy shortages

The ICC and random intercept variance for month were zero implying that the probability of delivering in an institution did not vary across months. Month was therefore removed from the model, reducing the levels to three. The data for the multilevel modelling had a level one sample size of 39,220 deliveries nested within 25 states, which is in turn nested within 2 rounds of data collection in almost each state, making the number of groups 41. The within state sample size ranged from 163 to 4,931 with an average state sample size of 1,568.8. The within round sample size 122 and 4,492 with an average of 956 deliveries in a round. The ICC for states was 24 percent, and that at the round level was 11.2 percent. The ICC for the level of states nested within round was 35 percent. Thus, 24 percent of the variation in the chances of delivering in an institution is due to state differences, while 11.2 percent is accounted for at the round level, and 35 percent of variation in the chances of delivering in an institution is accounted at the level of states nested within rounds. The test statistic of the likelihood ratio test was 13,192 with a p-value of 0.00 rejecting the null hypothesis that the conventional regression is better than multilevel modelling. The variances of *u*_0*s*_ and ω_0*t*_ were 0.509 and 0.483 respectively for states and round in the logistic regression and 0.842 and 0.143 respectively for multinomial regression. The variance indicates variations in institutional deliveries across states and rounds for data collection. All provide a strong justification for the use of multilevel modelling rather than the conventional regression.

Table 2 describes the prediction of institutional deliveries by energy shortages using both multilevel logistic and multinomial regressions. In general, energy shortages were linked to a reduced likelihood of institutional delivery; The results of the multilevel binary logistic regression demonstrate that a 1% increase in energy shortage was associated with about 1.0% (OR 0.993) lower odds of delivering in a healthcare institution, controlling for covariates. Similarly, the three-level multinomial regression revealed that, compared to delivering at home, the odds of delivering in a public healthcare facility and private healthcare facility were 1% (OR 0.992) and 0.3% (OR 0.997) lower, respectively, for each 1% increase in energy shortage, holding all other covariates constant. All findings were statistically significant at the 1% level.

**Table 2:** Effects of power shortages on institutional deliveries.

| Model | Logit Odd Ratios<br>Dummy<br>Institutional<br>Delivery | Multinomial Logit Odd Ratios<br>Base outcome: non-institutional<br>delivery |  | Multinomial Logit Odd Ratios<br>Marginal Predictions |  |  |
| --- | --- | --- | --- | --- | --- | --- |
|  |  | Public Facility | Private Facility | Non-Institutional | Public Facility | Private Facility |
| Energy Shortage (0-100) | 0.9929926***<br>(0.003268) | 0.9927166531***<br>(0.003336) | 0.99721102***<br>(0.003452) | 0.0011597<br>(0.0004744) | -0.0009394<br>(0.0003798) | -0.0002204<br>(0.0002966) |
| Mother's age at delivery (base: 15-19) |  |  |  |  |  |  |
| 20-24 | 0.8785811**<br>(0.047546) | 0.906486356***<br>(0.059889) | 0.82516748***<br>(0.071720) | 0.0182135<br>(0.0074169) | -0.0023689<br>(0.0083099) | -0.0158446<br>(0.007642) |
| 25-29 | 0.8074149**<br>(0.044749) | 0.821229558***<br>(0.061243) | 0.776300415***<br>(0.072793) | 0.028679<br>0.0631342 | -0.0792196<br>0.0670807 | 0.0505406<br>0.0533058 |
| 30-34 | 0.8519025***<br>(0.052212) | 0.874753<br>(0.067397) | 0.80742613***<br>(0.079512) | 0.0297962<br>(0.0076361) | -0.01327<br>(0.0084286) | -0.0165262<br>(0.0076754) |
| 35-39 | 0.910091<br>(0.069357) | 0.896323<br>(0.083926) | 0.923772<br>(0.097539) | 0.0134284<br>(0.010441) | -0.0113302<br>(0.0112851) | -0.0020982<br>(0.0100491) |
| 40-44 | 0.823851<br>(0.101223) | 0.846533<br>(0.134582) | 0.782819<br>(0.163631) | 0.0267672<br>(0.0168087) | -0.0092515<br>(0.0185504) | -0.0175157<br>(0.0172375) |
| 45-49 | 0.4672847***<br>(0.134073) | 0.446588719***<br>(0.331011) | 0.487683<br>(0.388482) | 0.1057338<br>(0.0396307) | -0.0748093<br>(0.0481528) | -0.0309245<br>(0.04303) |
| Education (base: no education) |  |  |  |  |  |  |
| Primary | 1.349012***<br>(0.057444) | 1.52016883***<br>(0.046971) | 1.058607<br>(0.060446) | -0.0388248<br>(0.0061178) | 0.0592232<br>(0.0083214) | -0.0203984<br>(0.0072526) |
| Secondary | 2.081864***<br>(0.079045) | 2.12339367***<br>(0.042343) | 1.988986<br>(0.049495) | -0.0996268<br>(0.0062665) | 0.0687971<br>(0.0069549) | 0.0308297<br>(0.0052035) |
| Higher | 5.568766**<br>(0.482294) | 3.46258008***<br>(0.095032) | 7.393535<br>(0.093169) | -0.2084756<br>(0.013466) | 0.0576826<br>(0.0107871) | 0.150793<br>(0.0099371) |
| Wealth Quintiles (base: poorest) |  |  |  |  |  |  |
| Q2 | 1.494261**<br>(0.120804) | 1.428228692***<br>(0.064942) | 1.67963968***<br>(0.099240) | -0.0569975<br>(0.0083034) | 0.0201312<br>(0.0107385) | 0.0368663<br>(0.0116015) |
| Q3 | 2.146809***<br>(0.188217) | 2.003256086***<br>(0.062418) | 2.56618304***<br>(0.093304) | -0.1076152<br>(0.0085134) | 0.0436453<br>(0.0101303) | 0.0639699<br>(0.0109287) |
| Q4 | 3.252794***<br>(0.188217) | 2.70865932***<br>(0.064297) | 4.92842763***<br>(0.092418) | -0.1667459<br>(0.009374) | 0.0469249<br>(0.0101637) | 0.119821<br>(0.0117316) |
| Richest | 5.646153***<br>(0.377025) | 3.30392778***<br>(0.074277) | 12.8073856***<br>(0.098236) | -0.2324501<br>(0.0109652) | 0.0152631<br>(0.0125667) | 0.217187<br>(0.0158917) |
| N | 77,583 | 77,583 | 77,583 | 77,583 | 77,583 | 77,583 |
| Robust standard errors in parentheses |  |  |  |  |  |  |
| *** p<0.01, ** p<0.05, * p<0.1 |  |  |  |  |  |  |

Notes: Additional controls for urbanicity, region, ethnicity of the mother, month of birth, and WHS round were also included in the regressions.

Marginal predictions were also computed for the multinomial logit specification. The results, shown in Table 2, columns 5-7, describe the increase or decrease in the likelihood of delivering at home or in a public or private facility due to energy shortages, evaluated at the mean of all other control variables. Thus, a 1% increase in energy shortage corresponds with a 0.12% increase in the likelihood of giving birth at home, and a 0.1% and 0.022% reduction in the likelihood of giving birth in a public and private healthcare institution, respectively. All marginal predictions were statistically significant.

The marginal predictions of a ten percent increase in energy shortage on the probability of institutional delivery were also calculated and are displayed in Table 3. Overall, a 10% increase in energy shortage was found to be associated with a 1.1% increase in the likelihood of giving birth at home, and a 0.01% and 0.002% reduction in the likelihood of giving birth in a public and private healthcare institution, respectively. All predictions were statistically significant. We also tested whether or not rain might be a potential confounder in our estimates. The results from those regressions, similar to those presented in Table 2, are presented in Appendix Table 1. Rain did not have any appreciable effect on institutional deliveries and therefore our results after controlling for rain are unchanged.

**Table 3:** Interactions power shortages and soci economic status.

| Model | Logit Odd Ratios | Multinomial Logit Odd Ratios |  |
| --- | --- | --- | --- |
|  | Dummy Institutional Delivery | Base outcome: non-institutional delivery<br>Public Facility | Private Facility |
| Energy Shortage X Wealth Quintiles (base: poorest) |  |  |  |
| Q2 | 0.998544<br>(0.005511) | 0.99614226<br>(0.0062305) | 1.003878502<br>(0.0094754) |
| Q3 | 1.003069<br>(0.0052046) | 1.002193803<br>(0.0058161) | 1.004332056<br>(0.008815) |
| Q4 | 1.005191<br>(0.0052892) | 1.005930315<br>(0.0059087) | 1.002839123<br>(0.0086675) |
| Richest | 1.005248<br>(0.0059741) | 1.011254662<br>(0.006679) | 0.996826745<br>(0.0091156) |
| Energy Shortage X Urban | 1.006138***<br>(0.0029826) | 1.003094378<br>(0.0033082) | 1.01092666***<br>(0.0036978) |
| Urban | 1.872921***<br>(0.1003322) | 2.07091450***<br>(0.0580503) | 1.575369109***<br>(0.0652458) |
| N | 77,583 | 77,583 | 77,583 |

Robust standard errors in parentheses
\*\*\* p<0.01, \*\* p<0.05, \* p<0.1
**Notes:** Additional controls used in Table2 and for urbanicity, region, ethnicity of the mother, month of birth, and WHS round were also included in the regression.

### Socioeconomic and urbanicity differences in the effect of energy shortages on institutional delivery

To assess whether wealth quintile or urbanicity modify the effect of energy shortages on institutional delivery, we added two interaction terms to the logistic and multinomial regressions described above, the results of which are presented in Table 4. Overall, we found little evidence that education and wealth quintile modifies the strength of the association between energy shortages and institutional delivery in either specification. Conversely, a statistically significant interaction between energy shortages and urbanicity was observed for private healthcare facilities in the multinomial regression, suggesting that rural mothers are less likely than urban mothers to give birth in a private health facility as energy shortages increase.

**Table 4:** Marginal effects of10%increase in energy shortage.

| Facility of delivery: | Original | 10% Increase | Change |
| --- | --- | --- | --- |
| Non-Institutional | 0.105 | 0.116 | 0.011*** |
| Public | 0.094 | 0.085 | -0.009*** |
| Private | 0.022 | 0.02 | -0.002*** |

\*\*\* p<0.01, \*\* p<0.05, \* p<0.1
Notes: The marginal effects of a 10% increase in energy shortage were evaluated for the multinomial logit of Table 2. The column "change" represents the expected increase (or decrease) in the delivery rates on each facility.

## Discussion

This study evaluated the association between energy shortages and institutional delivery rates in India using data from the 1998-99 and 2005-06 India DHS. Our results demonstrated that access to electricity was a significant predictor of institutional delivery, and a 10% increase in energy shortage was associated with 1.1% probability of giving birth outside a healthcare facility, holding all other factors constant. Given that there were close to 27 million births in India in 2011 and the institutional delivery rate was 78.5%, this translates to an additional 63,811 home births for each 10% increase in energy shortages.

These findings add to the growing body of literature exploring the effects of unreliable electricity on maternal healthcare utilization and support the results of previous studies in India, such as those by Köroglu et al. (23), who identified a significant reduction in the likelihood of delivering in an institution as power outage frequency increased, and Ghosh (35), who observed a significant association between a district development index, which included electrification, and a maternal healthcare index, which included safe delivery practices. Contrary to many recent studies that evaluated the effects of power outages occurring specifically in healthcare facilities on maternal outcomes, including those by Mbonye et al. (9), Erim et al. (30), Desalegn et al. (36), Singh et al. (33), and Bhattacharya et al. (29) who all found that poor electricity in health institutions reduced their capacity to provide high quality maternity care and emergency obstetric care, our findings revealed that energy shortages need not occur only in health facilities to have a significant influence on maternal care-seeking behaviours. Energy shortages in the region of residence, after all, limit access to healthcare institutions, for example by disrupting transportation and communication networks, which may also prevent women from giving birth in a facility.

Along with reduced odds of institutional delivery, our findings also revealed that the effect of energy shortages on institutional delivery varied according to the type of healthcare facility in question, and a greater reduction in the odds of giving birth in a facility was observed for public hospitals compared to private: 1.0% versus 0.3%. Interestingly, this is in contrast to what was observed in previous work by Köroglu et al. (23), who found that increased power outage frequency lowered the odds of delivering in a private healthcare facility to a greater degree than in a public healthcare facility. This could reflect the fact that private institutions have more resources at their disposal. Private healthcare institutions may be more likely to have backup generators, for example, which allow them to continue to provide patient care even during energy shortages. Alternatively, the greater impact of energy shortages on delivery in public institutions may be because private institutions tend to be concentrated in urban areas, which are easier to access during times of power outage, or because the people who seek care at private facilities are typically of higher socioeconomic status, and may therefore have access to alternative means of transportation and communication. Indeed there is substantial evidence to suggest that private healthcare institutions overwhelmingly serve urban areas and the rich (37), and that poorer women and women who live further from a health facility are less likely to have an institutional birth (38,39).

To explore the possible moderating prediction of income and urbanicity in greater detail we conducted an interaction analysis in both logistic and multinomial regressions. While education and wealth quintile were not found to moderate the link between energy shortages and institutional delivery in either specification, the interaction term for energy shortage and urbanicity was statistically significant in the multinomial regression for private facilities only, suggesting that a rural woman’s odds of delivering in a private institution are more sensitive than a public institution in times of energy shortage, compared to an urban woman. This is contrary to Koroglu, et al., (23) which examined effect of power outage on institutional deliveries in the Indian state Maharashtra and found that increased frequencies in power outages was associated with lower institutional deliveries in private facilities for low-wealth women as well as in public facilities for rural women. The results in the current study implies that at the national level such effects are not statistically significant but there could be state variation. The higher response of rural women who deliver in private facilities may be because private facilities are more likely to be located in urban areas and are therefore less accessible to rural women during power outages, who are unable to travel the long distance required to visit these institutions.

## Implications

The results of this study illustrate the importance of reliable electricity for maternal healthcare utilization and provide new evidence of a significant negative association between energy shortages and institutional delivery rates in India. Access to consistent and reliable electricity remains an important policy issue in India, and power outages frequently cause disruptions, not only to health facilities’ operations, but to care seeking behaviours as well. Given that institutional delivery is an important predictor of maternal mortality, efforts must be made to mitigate the effects of energy shortages on maternal healthcare seeking and utilization. Such strategies could include expanding access to alternative sources of power at healthcare facilities, for example by mandating backup generators. Indeed, as Koroglu et al. (23) showed in their recent study, more than half of health facilities (53%) in Maharashtra State, India did not have a generator in 2011, while over three quarters (78%) reported having no electricity or experiencing a power outage almost every day. Ensuring that all healthcare institutions have a backup power source would therefore allow them to maintain basic operations even during an energy shortage, which would enable institutional delivery. Additionally, programs and policies that ensure access to care is maintained during an energy shortage should be implemented, particularly in rural communities. Expanded telehealth and midwife services, for example, could allow women who are not able to travel to a health facility for any reason, including a power outage, to receive skilled care, lowering the likelihood of maternal and infant mortality (40). Finally, and most importantly, resources must be invested to improve the quality of the energy supply in India, such that the frequency of energy shortages is reduced. Recent efforts to improve electricity in India have shown some success, most notably Prime Minister Narendra Modi’s achievement at electrifying all villages in April 2018 (19), however electricity reliability continues to be an issue and many individual homes lack a grid connection, with rural women the most disadvantaged.

## Limitations

The study relies upon self-reported data collected from households on births and deliveries, and may therefore have been vulnerable to various types of survey bias, including recollection bias or social desirability bias. Second, the data in question is outdated, collected between 15 and 25 years ago, and the state of both electricity and institutional delivery in the country have likely changed in the decades since the data was collected. Given that this energy data is unique, however, compiled by Allcott et al. in conjunction with India’s Central Electricity Authority (1), and that more recent data is not yet available, we feel the benefits of its use outweigh any potential risks. Third, the study did not include possible energy management policies such as availability of backup energy in case of black out. The presence of such policies could reduce the association between power outages and institutional deliveries.

## Conclusions

This study explored the association between energy shortages in the state of residence and institutional delivery in India. Our findings revealed that energy shortages were a significant predictor of a reduced odds of institutional delivery, and that public institutions were affected to a greater degree than private institutions. Given that institutional delivery is considered a protective factor for a number of adverse maternal and infant health outcomes, including death, there is an urgent need for government to improve the reliability of electricity in India, and to develop strategies to ensure women are able to access maternal care, including institutional delivery, even during energy shortages.

## Data Availability

Data used are all in public repositories. The Indian Demographic Health Surveys 1998/9 to 2005/6. The data on electricity can be found at www.indiaenergydata.info

https://dhsprogram.com/methodology/survey/survey-display-156.cfm

## Acknowledgements

We acknowledge that this work was conducted on the Haldimand Tract, traditional territory of the Neutral, Anishnaabe and Haudenosaunee peoples. We are grateful to the AXA Research Foundation for financial support for this project.

---

**Appendix Table 3:** Effects of power shortages on Institutional deliveries - with rain control.

| Model | Logit Odd Ratios<br>Dummy<br>Institutional<br>Delivery | Multinomial Logit Odd Ratios<br>Base outcome: non-institutional<br>delivery |  |
| --- | --- | --- | --- |
|  |  | Public Facility | Private Facility |
| Energy Shortage (0-100) | 0.998***<br>(0.00327) | 0.995***<br>(0.00334) | 0.991***<br>(0.00345) |
| Rain at month of delivery (mm) | 1<br>0 | 1<br>0 | 1<br>0 |
| Mother's age at delivery (base: 15-19) |  |  |  |
| 20-24 | 0.8785811**<br>(0.047546) | 0.906486356***<br>(0.059889) | 0.82516748***<br>(0.071720) |
| 25-29 | 0.8074149**<br>(0.044749) | 0.821229558***<br>(0.061243) | 0.776300415***<br>(0.072793) |
| 30-34 | 0.8519025***<br>(0.052212) | 0.874753<br>(0.067397) | 0.80742613***<br>(0.079512) |
| 40-44 | 0.910091<br>(0.069357) | 0.896323<br>(0.083926) | 0.923772<br>(0.097539) |
| 45-49 | 0.823851<br>(0.101223) | 0.846533<br>(0.134582) | 0.782819<br>(0.163631) |
|  | 0.4672847***<br>(0.134073) | 0.446588719***<br>(0.331011) | 0.487683<br>(0.388482) |
| Education (base: no education) |  |  |  |
| Primary | 1.349012***<br>(0.057444) | 1.52016883***<br>(0.046971) | 1.058607<br>(0.060446) |
| Secondary | 2.081864***<br>(0.079045) | 2.12339367***<br>(0.042343) | 1.988986<br>(0.049495) |
| Higher | 5.568766**<br>(0.482294) | 3.46258008***<br>(0.095032) | 7.393535<br>(0.093169) |
| Wealth Quintiles (base: poorest) |  |  |  |
| Q2 | 1.494261**<br>(0.120804) | 1.428228692***<br>(0.064942) | 1.67963968***<br>(0.099240) |
| Q3 | 2.146809***<br>(0.188217) | 2.003256086***<br>(0.062418) | 2.56618304***<br>(0.093304) |
| Q4 | 3.252794***<br>(0.188217) | 2.70865932***<br>(0.064297) | 4.92842763***<br>(0.092418) |
| Richest | 5.646153***<br>(0.377025) | 3.30392778***<br>(0.074277) | 12.8073856***<br>(0.098236) |
| N | 69,330 | 69,330 | 69,330 |
| Robust standard errors in parentheses |  |  |  |
| *** p<0.01, ** p<0.05, * p<0.1 |  |  |  |

**Notes:** Additional controls for urbanicity, region, ethnicity of the mother, month of birth, and WHS round were also included in the regressions.

## Footnotes

2 https://data.gov.in/resources/sub-divisional-monthly-rainfall-1901-2017

